# The Impact of Group II Pulmonary Hypertension on Congestive Heart Failure Patients Admitted with ST Elevation Myocardial Infarction, A Nationwide Study

**DOI:** 10.1101/2023.11.21.23298866

**Authors:** Mohamad El Labban, Mikael R Mir, Alexandra Abruzzo, Sydney Boike, Fayreal A Niaz, Natasha T Vo, Ibtisam Rauf, Syed Anjum Khan

## Abstract

**Objective:** To study the impact of group II pulmonary hypertension (PH) on the outcomes of patients admitted with ST-elevation Myocardial Infarction (STEMI), we conducted a nationwide retrospective cohort study.

**Patients and Methods:** Using the National Inpatient Sample (NIS) Database from 2017 to 2020, a retrospective study of adult patients with a principal diagnosis of STEMI with a secondary diagnosis with or without group II PH according to ICD-10 codes. Several demographics, including age, race, and gender, were analyzed. The primary endpoint was mortality, while the secondary endpoints included cardiogenic shock, mechanical intubation, length of stay in days, and patient charge in dollars. Multivariate logistic regression model analysis was used to adjust for confounders, with a p-value less than 0.05 considered statistically significant.

**Results:** The study included 27,020 patients admitted with a STEMI, 95 of whom had group II PH. The mean age for patients with and without PH was 66 and 67, respectively. In the PH group, 37% were females compared to 34% in the non-PH group. The in-hospital mortality rate was higher in the PH group (31.5% vs. 9.5%, *P <.001*, aOR 3.25, *P <.023*). The rates and adjusted odds of cardiogenic shock and mechanical ventilation were higher in the PH groups (aOR 1.12 aOR 2.16, respectively) but not statistically significant. Patients with PH had a longer length of stay and a higher total charge.

**Conclusion:** Group II PH was associated with worse clinical and economic outcomes in heart failure patients admitted with STEMI.

## Introduction

Coronary artery disease (CAD) is the leading cause of death, disability, and disability-adjusted life years worldwide, and its prevalence continues to climb[1, 2]. Myocardial infarction (MI) occurs due to coronary artery disease and is classified as transmural MI and non-transmural MI. In contrast to a non-transmural MI, a transmural MI affects the epicardium, myocardium, and endocardium[3]. ST segment elevations or depressions are key findings that allow an MI to be detectable on electrocardiograms (ECGs). Patients with presenting symptoms are streamlined into different treatment regimens based on the ST elevation (STEMI) versus non-ST elevation (NSTEMI) paradigm. This paradigm can delay life-saving treatments, i.e., emergent reperfusion, in patients without suspecting ECG findings yet with complete acute coronary occlusion [4].

Pulmonary hypertension (PH) is characterized by elevated blood pressure in the pulmonary arteries, resulting in increased blood flow resistance and reduced oxygen delivered to the body’s tissues. PH is defined as a mean pulmonary arterial pressure (PAP) ≥ 20 mmHg at rest, measured via right heart catheterization [5]. However, a clinical diagnosis can be made with significant left heart disease or chronic lung disease. The pathogenesis of PH is multifactorial and can involve multiple systems; therefore, PH is often subdivided into five groups based on pathophysiology (Table 1). The link between PH and MI has been established, with PH being a known risk factor for MI through right ventricular hypertrophy, decreased cardiac output, and subsequent ischemia of the myocardium, in addition to other mechanisms. In this study, we further explore the clinical and economic impact of Group 2 PH (G2PH) on patients admitted with STEMI.

**Table 1.** Clinical classification of pulmonary hypertension (PH) [6].

| Group | Characteristics |
| --- | --- |
| Group 1: Pulmonary arterial hypertension | 1.1 Idiopathic PAH<br>1.2 Heritable PAH<br>1.3 Drug- and toxin-induced PAH<br>1.4 PAH associated with connective tissue disease, HIV, portal hypertension, congenital heart disease, Schistosomiasis<br>1.5 PAH long-term responders to calcium channel blockers<br>1.6 PAH with over features of venous/capillaries involvement<br>1.7 Persistent PH of the newborn syndrome |
| Group 2: PH due to left heart disease | 2.1 PH due to heart failure with preserved LVEF<br>2.2 PH due to heart failure with reduced LVEF<br>2.3 Valvular heart disease<br>2.4 Congenital/acquired cardiovascular conditions leading to post-capillary PH |
| Group 3: PH due to chronic lung disease and/or hypoxemia | 3.1 Obstructive lung disease<br>3.2 Restrictive lung disease<br>3.3 Other lung disease with mixed restrictive/obstructive pattern |
|  | 3.4 Hypoxia without lung disease<br>3.5 Developmental lung disorders |
| Group 4: PH due to pulmonary artery obstructions | 4.1 Chronic thromboembolic PH<br>4.2 Other pulmonary artery obstructions |
| Group 5: PH due to multifactorial mechanisms | 5.1 Hematologic disorders<br>5.2 Systemic and metabolic disorders<br>5.3 Others<br>5.4 Complex congenital heart disease |

## Materials and Methods

### Design and description of the database

We conducted a retrospective cohort study using the national inpatient sample (NIS) from 2017 to 2020. The NIS is part of the Healthcare Cost and Utilization Project (HCUP) and is sponsored by the Agency for Healthcare Research and Quality (AHRQ) [7]. The NIS is the largest inpatient hospital discharge database in the United States. Hospitals are stratified according to ownership/control, bed size, teaching status, urban/rural location, and geographic region. A 20% probability sample is collected from all hospitals within each stratum. All discharges from these hospitals are recorded and weighted to ensure national representation.

### Data user agreement

Dr. El-Labban (first author) completed the data user agreement with HCUP-AHRQ. The HCUP datasets are publicly available and hence are considered exempt from full or expedited institutional review boards (IRB) review (Federal Regulations 45 CFR 46.101 (b).

### Selection of cases and outcome variables examined

In the NIS dataset, the principal diagnosis is the main ICD-10 code of the hospitalization. It does not have to be the reason for admission to the hospital. All procedure codes detected via NIS are linked to the hospitalization. In our study, STEMI was selected as the principal diagnosis (International Classification of Disease, 10th edition, clinical modification [ICD-10-CM]). Our inclusion criteria included adult patients (age 18 years or older) with a history of congestive heart failure presenting with a non-elective/urgent admission under a principal diagnosis of STEMI from 2017 to 2020. We then divided the sample into a group with associated group II pulmonary hypertension vs. control (those without group II pulmonary hypertension). ICD-10 codes were also used to identify secondary diagnoses that included diabetes mellitus (DM), essential hypertension, supraventricular tachycardia (SVT), chronic heart failure (CHF), chronic obstructive pulmonary disease (COPD), chronic kidney disease (CKD), dementia, obesity, and sepsis. The Charlson Comorbidity Index (CCI) also described patients’ comorbidities. Outcomes, including mortality, cardiogenic shock, mechanical ventilation, length of stay, and total charges, were generated from the NIS dataset. We excluded patients with acute exacerbation of heart failure and pulmonary hypertension other than group II.

### Statistical Analysis

Statistical analyses were conducted using STATA BE Version 17.0. All statistical tests were two-sided, and a p-value of <0.05 was considered statistically significant. Chi-square analysis was used to describe the difference in patients’ characteristics and secondary diagnoses according to the presence and absence of pulmonary hypertension. The impact of pulmonary hypertension on outcomes was defined using two methods: Chi-square analysis to compare outcomes’ rates and multivariable regression models to describe the isolated impact of pulmonary hypertension on the odds of the outcome.

## Results

Table 2 represents the patient demographics from the study and is separated based on the presence or absence of Group II PH. The average age of patients without PH was 67 years, and that of patients with PH was 66 years. The differences in the proportion of patients with and without PH across ethnicities were not statistically significant. CCI scores of three or more were relatively increased in the patients with G2PH compared to non-G2PH patients. All comorbidities were increased in patients with G2PH than in patients without G2PH; SVT AND CKD were substantially more prevalent in patients with PH. There were more than twice as many patients with SVT and CKD in the G2PH group.

**Table 2.**
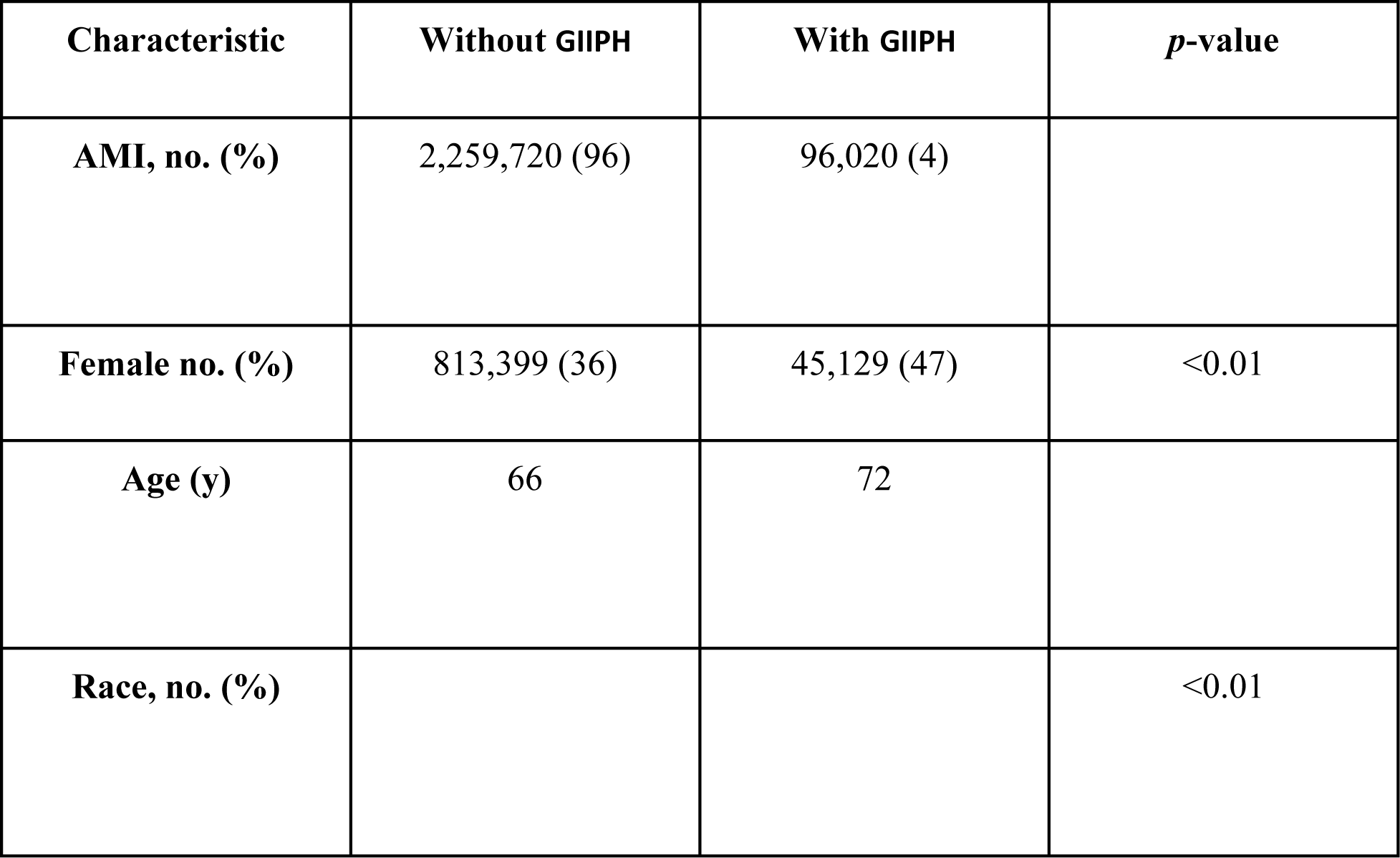
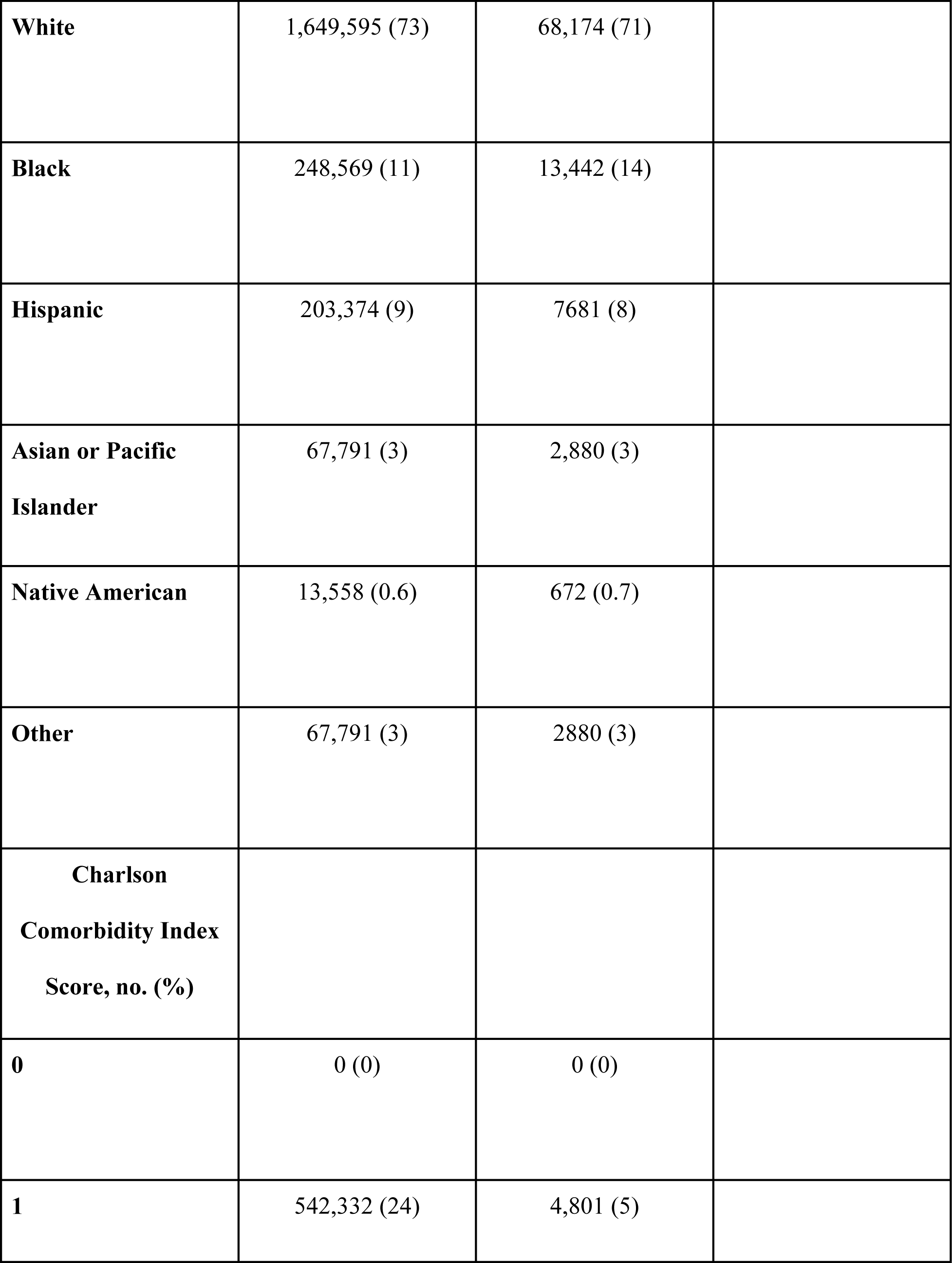
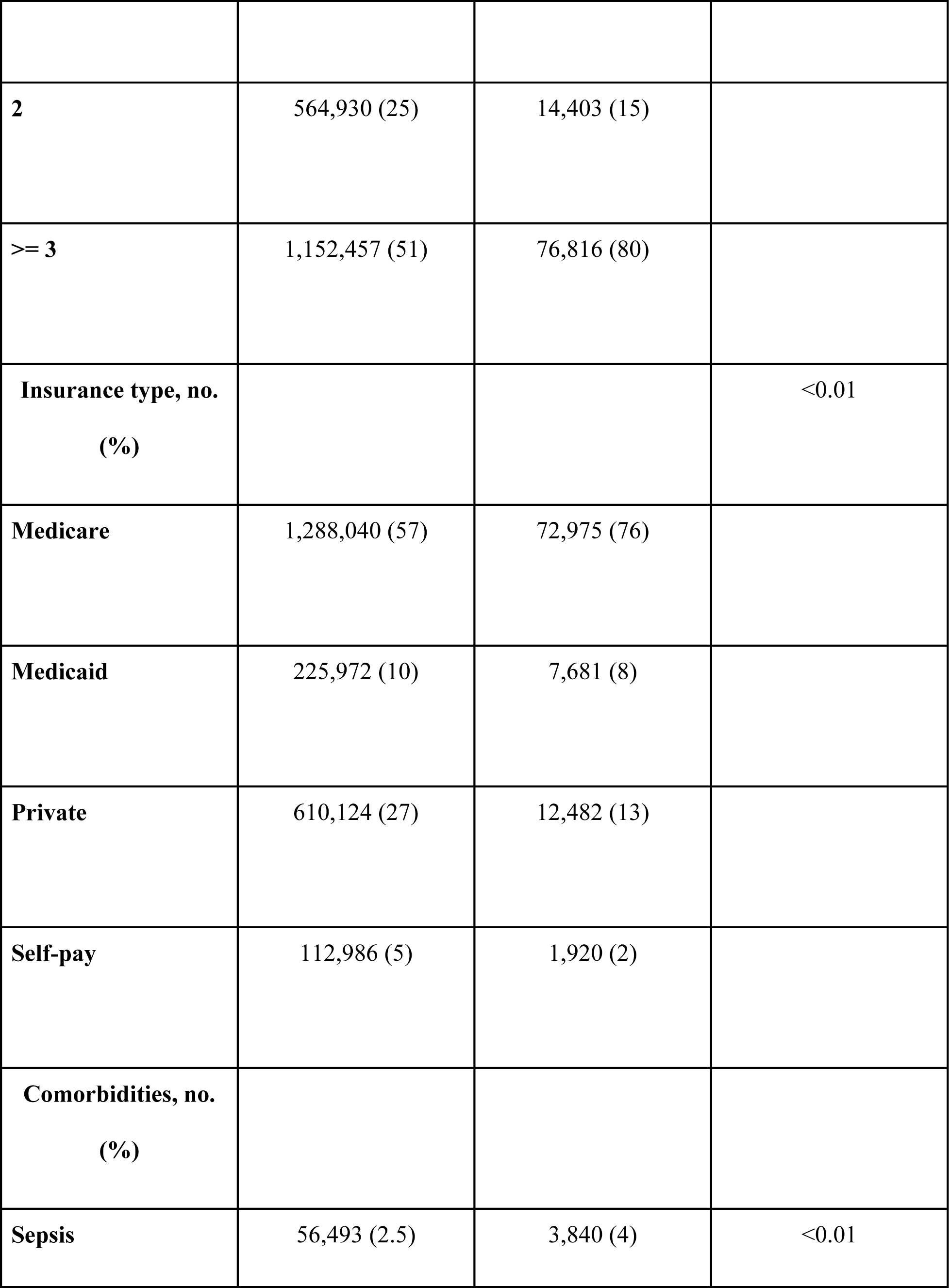
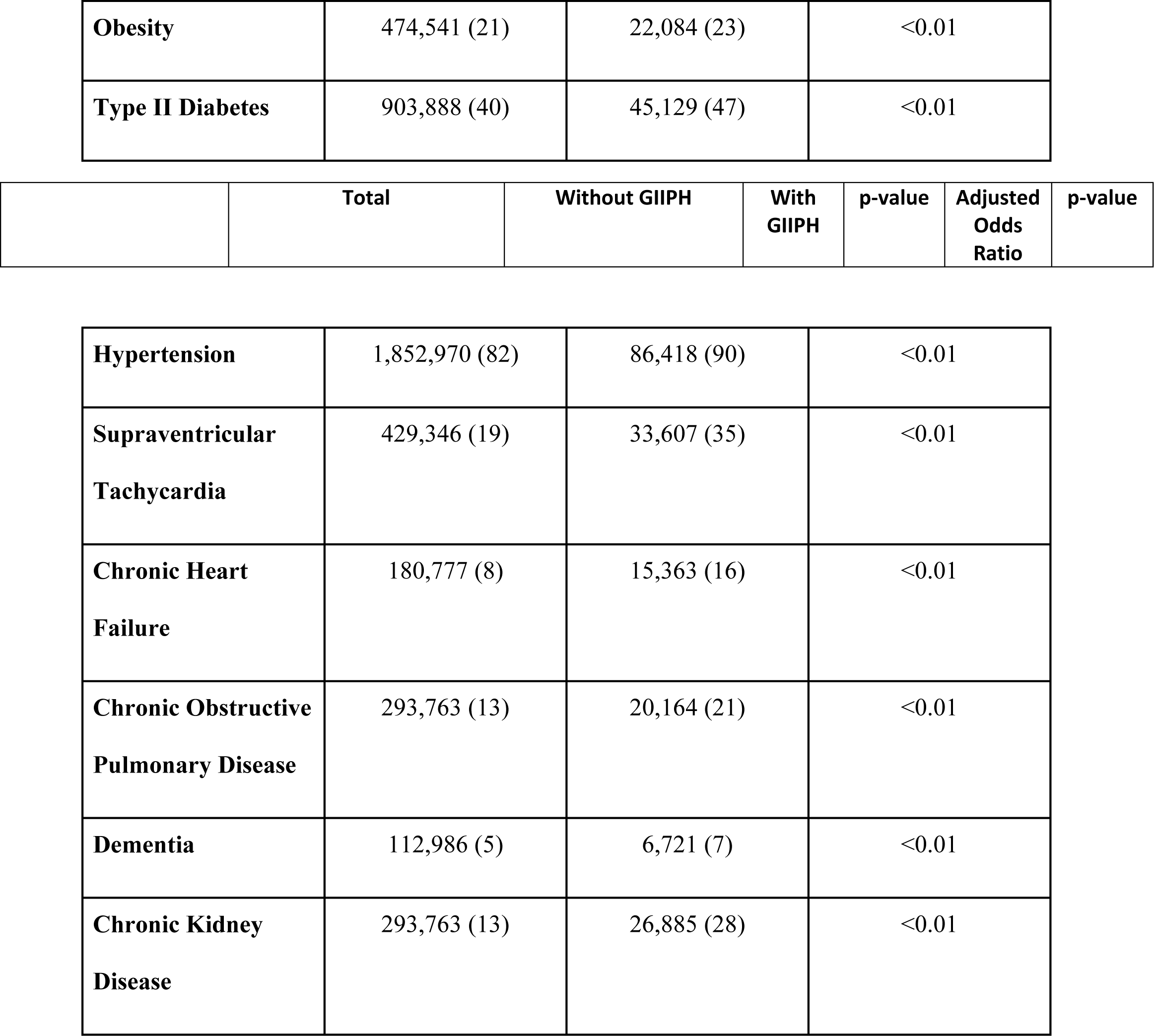
Patient Demographics.

| **Characteristic** | **Without GIIPH** | **With GIIPH** | ***p*-value** |
| --- | --- | --- | --- |
| **AMI, no. (%)** | 2,259,720 (96) | 96,020 (4) |  |
| **Female no. (%)** | 813,399 (36) | 45,129 (47) | <0.01 |
| **Age (y)** | 66 | 72 |  |
| **Race, no. (%)** |  |  | <0.01 |
| **White** | 1,649,595 (73) | 68,174 (71) |  |
| **Black** | 248,569 (11) | 13,442 (14) |  |
| **Hispanic** | 203,374 (9) | 7681 (8) |  |
| **Asian or Pacific**<br>**Islander** | 67,791 (3) | 2,880 (3) |  |
| **Native American** | 13,558 (0.6) | 672 (0.7) |  |
| **Other** | 67,791 (3) | 2880 (3) |  |
| **Charlson**<br>**Comorbidity Index**<br>**Score, no. (%)** |  |  |  |
| **0** | 0 (0) | 0 (0) |  |
| **1** | 542,332 (24) | 4,801 (5) |  |
| **2** | 564,930 (25) | 14,403 (15) |  |
| **>= 3** | 1,152,457 (51) | 76,816 (80) |  |
| **Insurance type, no.**<br>**(%)** |  |  | <0.01 |
| **Medicare** | 1,288,040 (57) | 72,975 (76) |  |
| **Medicaid** | 225,972 (10) | 7,681 (8) |  |
| **Private** | 610,124 (27) | 12,482 (13) |  |
| **Self-pay** | 112,986 (5) | 1,920 (2) |  |
| **Comorbidities, no.**<br>**(%)** |  |  |  |
| **Sepsis** | 56,493 (2.5) | 3,840 (4) | <0.01 |
| **Obesity** | 474,541 (21) | 22,084 (23) | <0.01 |
| **Type II Diabetes** | 903,888 (40) | 45,129 (47) | <0.01 |

|  | **Total** | **Without GIIPH** | **With GIIPH** | **p-value** | **Adjusted Odds Ratio** | **p-value** |
| --- | --- | --- | --- | --- | --- | --- |
| **Hypertension** | 1,852,970 (82) | 86,418 (90) | <0.01 |
| **Supraventricular Tachycardia** | 429,346 (19) | 33,607 (35) | <0.01 |
| **Chronic Heart Failure** | 180,777 (8) | 15,363 (16) | <0.01 |
| **Chronic Obstructive Pulmonary Disease** | 293,763 (13) | 20,164 (21) | <0.01 |
| **Dementia** | 112,986 (5) | 6,721 (7) | <0.01 |
| **Chronic Kidney Disease** | 293,763 (13) | 26,885 (28) | <0.01 |

Rates and odds of all-cause in-hospital mortality were significantly higher in patients with G2PH and STEMI (*p*<0.01) (Table 3). Patients with G2PH had worse secondary outcomes as well. Rates and odds of in-hospital cardiogenic shock and mechanical ventilation were relatively increased in patients with STEMI (Table 3).

**Table 3:**
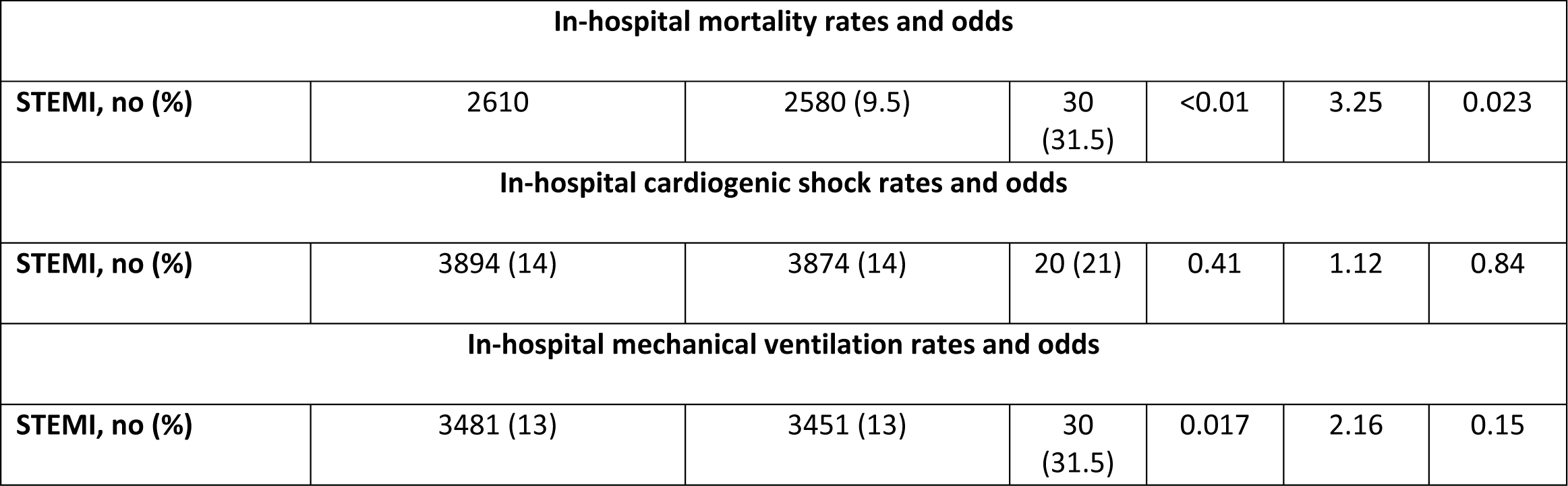
Primary and Secondary outcomes.

| In-hospital mortality rates and odds |  |  |  |  |  |  |
| --- | --- | --- | --- | --- | --- | --- |
| STEMI, no (%) | 2610 | 2580 (9.5) | 30<br>(31.5) | <0.01 | 3.25 | 0.023 |
| In-hospital cardiogenic shock rates and odds |  |  |  |  |  |  |
| STEMI, no (%) | 3894 (14) | 3874 (14) | 20 (21) | 0.41 | 1.12 | 0.84 |
| In-hospital mechanical ventilation rates and odds |  |  |  |  |  |  |
| STEMI, no (%) | 3481 (13) | 3451 (13) | 30<br>(31.5) | 0.017 | 2.16 | 0.15 |

Though not statistically significant, the mean length of stay and total hospital charges (Table 4) were more prolonged and higher in patients with PH and STEMI.

**Table 4:** Mean Length of Stay and Total Charges.

|  | Without GIIPH | With GIIPH |
| --- | --- | --- |
| Length of stay (Days) | 5 | 6.42 |
| Total charges (\$) | 125,708 | 150,439 |

## Discussion

### Demographics

Race had an effect on the presence of G2PH. Compared to the same race in the G2PH cohort, White patients had a higher proportion of patients without G2PH. [8] The discrepancy in the proportion of patients with and without Group II PH in one race may be due to extrinsic factors—for instance, lack of recruitment of minority races and socioeconomic status. Most research published on PH reflects White participants. Studies have shown that White patients had a significantly lesser degree of socioeconomic distress than their Black counterparts [9]. This is consistent with Black patients having a higher proportion of PH. Physician and medical research distrust among minority populations could account for discrepancies in race representation in research [10]. Additionally, there could be a lack of understanding of medical trials amongst minority races due to language barriers or the overuse of medical jargon.

### Individual comorbidities and the Charlson Comorbidity Index score

There were more patients in our analysis with CCI scores of three or more who had Group II PH. Given the relative increase in comorbidities, patients with Group II PH may generally be sicker. PH may be idiopathic but is also commonly associated with other diseases. Metabolic disorders, certain types of cancers, lung disease, and left heart disease are just a few diseases that put patients at risk for developing PH or are associated with PH [11]. There was a relative increase in the number of patients with each comorbidity with respect to the Group II PH cohort compared to the non-PH cohort. SVT and CKD were both significantly more prevalent in patients with G2PH compared to patients without PH. SVT can result in heart failure [12]. CHF can cause elevated hydrostatic pressures in the pulmonary vasculature, leading to the development of Group II PH. [13-15]. Similarly, there were significantly more patients with CKD in the PH cohort. CKD may incite pulmonary vascular dysfunction and eventual PH through the release of uremic toxins and subsequent inflammation and endothelial dysfunction; thus, group II PH is a highly prevalent disease in patients with CKD [16].

### In-hospital mortality cardiogenic shock

Patients with Group II PH had significantly higher in-hospital mortality and cardiogenic shock rates after aSTEMI compared to those without Group II PH, even after adjustment with the CCI. Although the in-hospital mortality outcome reflects all-cause inpatient mortality, those with group II PH have more systemic complications than those without group II PH, which could contribute to the increased risk of in-hospital mortality. Elevated PAP is associated with right ventricle (RV) afterload, myocardial dysfunction, and impaired hemodynamics. The impaired perfusion due to low cardiac output (CO) and the systemic congestion caused by impaired RV function affects multiple organs. For example, systemic consequences of PH include congestive hepatopathy in the liver, low perfusion renal injury in the kidneys, and increased systemic inflammation due to the release of soluble pro-inflammatory chemokines/cytokines [17]. Furthermore, elevated PAP in the chronic state also causes RV dysfunction that causes eccentric remodeling and contractile dysfunction of the RV [17]. The higher rates of cardiogenic shock are likely due to the RV dysfunction. RV dilation and an increase in RV size cause a mechanical septal leftward shift, leading to compression of the left ventricle (LV) [18]. As a result, the LV displays a “D-shape,” and there is an increased LV eccentricity index [17]. Furthermore, low cardiac output (CO) from the RV dysfunction can contribute to the underfilling of the LV, leading to reduced LV CO and further exacerbating the cardiogenic shock. This effect can be further exacerbated during MI, such as an inferior MI in which the RV is affected, further contributing to RV dysfunction. Cardiogenic shock is a severe complication of AMI; those with cardiogenic shock and STEMI have higher fatality rates than those with NSTEMI [19]. There was a relatively higher rate of cardiogenic shock in patients with PH/STEMI. This occurrence is likely due to ischemia that can occur as a result of chronic PH and RV dysfunction, as STEMI is due to transmural infarction and a greater burden of ischemia [19]. These consequences, in addition to RV failure and hemodynamic instability, contribute to the higher risk of in-hospital mortality and cardiogenic shock in these patients.

### Mechanical ventilation

Group II PH was an independent risk factor for needing mechanical ventilation after acute MI. There are no studies in the current literature that investigate this trend. One possible explanation is related to the effects of PH on skeletal muscles and the diaphragm. PH has been shown to adversely affect striated muscles other than the RV, such as the diaphragm and peripheral skeletal muscle [20]. Studies investigating the pathophysiology of this occurrence have yet to show consistent results. Still, some have shown evidence of both decreased maximal tension [21] and decreased slow twitch fibers in the diaphragm, which correlated with maximal inspiratory pressure.[20]

### Admission length of stay and total cost

Patients with Group II PH experienced longer hospital stays following MI after adjusting for comorbidities and their CCI scores. Patients had spent an average of 34.08 hours more in the hospital. One explanation for this is that after undergoing PCI, patients are at an increased risk of cardiomyocyte reperfusion injury that could result in heart failure [22]. These effects may be exacerbated in patients that already have Group II PH in addition to its associated right ventricular heart failure [23]. The likelihood of ventricular wall rupture and mortality are significantly increased in patients with elevated PAP [24]. Longer hospital stays, and consecutive post-MI interventions precipitate larger hospital costs, which are reflected in Table 4. Patients with Group II PH had more expensive hospital stays than patients without PH. In 2015, Plent et al. calculated the median cost for patients undergoing an uncomplicated PCI in their study to be about $18,419 [25]. Therefore, on top of the baseline cost of treating a patient with MI, patients with G2PH who are already at an increased risk of complications may end up costing the hospital hundreds to thousands of additional dollars.

### Limitations

There are some limitations to our study. We conducted a retrospective cohort study using the NIS, an administrative database that limits the uniformity of STEMI and G2PH diagnosis with potential misclassification secondary to the use of the ICD-10 CM codes. Second, because of the administrative nature of our database, our analysis did not include the severity of the disease at admission or the types, dosages, and frequencies of different STEMI-specific treatments. Third, the primary cause of mortality is not specified in NIS; therefore, we presented the total all-cause mortality. Finally, since this is not a randomized controlled trial, the conclusion that G2PH significantly impacts the outcomes of STMEI should be viewed with caution. Despite these limitations, this study is an important contribution to understanding the relationship between STEMI and Group II PH. This study is the first to investigate that relationship using the NIS and, hence, with a remarkably large sample size. Moreover, we investigated how G2PH affected the total charges during hospital stays, which is not traditionally included in most studies.

## Conclusion

Our study, investigating the clinical and economic impact Group II PH has on patients admitted to the hospital with an acute MI, reveals important insights into the clinical outcomes and management of this high-risk population. Our findings indicate that the presence of G2PH significantly impacts the prognosis of patients with acute MI. The demographic analysis highlights the need for tailored approaches, as certain populations may be more vulnerable to the detrimental effects of PH. Group II PH is an independent risk factor for in-hospital mortality, cardiogenic shock, and mechanical ventilation, underscoring the importance of early identification and appropriate management of this condition. These results emphasize the need for healthcare providers to be vigilant and proactive in recognizing and addressing PH in patients with STEMI to optimize patient outcomes and reduce morbidity and mortality.

Further research should focus on elucidating the underlying mechanisms behind why PH is an independent risk factor for post-MI complications and investigating targeted interventions to improve the prognosis of this complex patient population.

## Data Availability

A STATA log containing the manuscript's results is available upon request.

## List of abbreviations

AMI: acute myocardial infarction
CCI: Charlson comorbidity index
CKD: chronic kidney disease
DM: Diabetes mellitus
LV: left ventricle
PAP: Pulmonary arterial pressure
PH: pulmonary hypertension
MI: myocardial infarction
NSTEMI: non-ST-elevation myocardial infarction
RV: right ventricle
STEMI: ST-elevation myocardial infarction

## Acknowledgments

The authors report no potential competing interests.

## Author Contributions

Study concept and design—all authors; Data acquisition and statistical analyses—Dr. El Labban; Interpretation of data—Dr. El Labban; Drafting of manuscript—All authors; Critical revision of the manuscript for important intellectual content—all authors.

## Notes

### Competing Interest Statement

The authors have declared no competing interest.

### Clinical Trial

n/a

### Funding Statement

no external funding was received.

### Author Declarations

The HCUP datasets are publicly available and hence are considered exempt from full or expedited institutional review boards (IRB) review (Federal Regulations 45 CFR 46.101 (b).

